# Estimation of uncertainty in Loa loa microfilarial load by microscopy

**DOI:** 10.1101/2023.02.19.23286133

**Authors:** Talagbé Gabin Akpo

## Abstract

For determine the uncertainty of reading and measurement, as well as the Pari intervals of microfilarial load or microfilaremia (mf) per millimeter of Loa loa performed by microscopy. It is important to consider the uncertainty in the measurement or reading of the Loa Loa microfilarial load for the administration of ivermectin.

We review existing methods for calculating the uncertainty in the measurement of a particular quantity, with emphasis on the one proposed in GUM. The data used here come from research conducted by CRFilMT in Ebolowa and Mbalmayo in 2007 and 2010, respectively, and in the Okola health district in Cameroon in 2015. The data consist of several measurements or readings of Loa loa load on each sampled individual. The application of the GUM method to our data was done using a 2-level hierarchical model.

We estimated the uncertainty and sources of variation in the measurements and readings of Loa loa microfilarial load, and provided 95% intervals for the true values (8,000 mf/mL and 30,000 mf/mL), of this load for each individual. For reading, the reading uncertainty is 3.84 with a Pari interval of [6, 723.15, 11, 264] of the 8,000 mf/mL microfilar charge and 7.45 with a Pari interval of [26, 819.55, 35, 152.09] of the 30,000 mf/mL microfilar charge. For the measurement, the reading uncertainty is 20.93 with a Pari interval of [7, 647.32, 8, 216.26] of the 8,000 mf/mL microfilar charge and 40.53 with a Pari interval of [26, 819.55, 35, 152.09] of the 30,000 mf/mL microfilar charge.

## Introduction

Loa loa is the parasite that causes the condition known as “loasis”. It is transmitted to humans by tabanidae of the genus Chrysops, the principal vector species of which are C. silacae and C. dimidiata, Duke (1994). It is endemic in tropical, equatorial forest and savanna regions of West, Central and East Africa, Kershaw and al. (1953); Fain (1978)). It is one of the main causes of medical consultation in the areas where it is endemic, because of the nuisance caused (itching, pruritus, oedema, etc.). However, it is still considered a benign condition. Loiasis has received renewed attention in recent years because of the serious neurological and sometimes fatal side effects (Boussinesq & Chippaux (2003) ; Kamgno and al. (2016)) that have occurred following treatment of onchocerciasis or lymphatic filariasis with ivermectin, see Ottesen (2006), in areas where loiasis is coendemic. These side effects usually occur in people with high loasis loads. Indeed, who recommends that hypo-endemic areas for onchocerciasis and endemic areas for loasis, or hypo-endemic areas for lymphatic filariasis and endemic areas for loasis, should not be treated because in these areas the risk of side effects is greater than the benefit of treatment at the community level. These areas excluded from treatment may form foci of loasis recontamination, thus constituting an obstacle to disease elimination, Zoure and al. (2011). Several alternative treatment strategies in these loasis and lymphatic filariasis or loasis and onchocerciasis co-endemic areas are possible, including the strategy of early identification of individuals at risk of side effects and excluding them from mass ivermectin treatment, Fobi and al. (2000). To identify individuals who may develop serious side effects, risk thresholds were defined in relation to Loa loa microfilarial load. People with microfilaria loads between 8,000 and 30,000 microfilaria/ml receive the treatment, but regular follow-ups are made by medical teams are done by medical teams. However, those with microfilaria loads above 30,000 mf/ml are at risk of serious, sometimes fatal, side effects and are excluded from taking ivermectin.

## 1 Materials used

The data used in this study were collected during several population-based research projects conducted by the Centre de Recherche sur les Filarioses et les autres maladies tropicales (CRFilMT) with the aim of eliminating lymphatic and meningeal filariasis in Cameroon, or loase, which is prevalent in Central Africa, specifically in Cameroon. Indeed, Cameroon is an endemic country for loasis. Faced with this situation, vast programs for the elimination of loasis have been developed throughout the national territory. This work has led to the clarification of important research questions in several areas. The endemic areas covered by these programs are shown on the map in 1.

The first database (Base 1) comes from population-based research conducted by CR-FilMT in two cities in southern and central Cameroon: Ebolowa in 2007 and Mbalmayo in 2010. The survey collected socio-demographic data (sex, age and place of residence) of the population of these two cities, but also enumerated the microfilarial density of Loa loa per thick drop of calibrated blood. The data collected in this database, as mentioned above, will allow us to estimate the uncertainty in the measurement of the microfilarial load of Loa loa. The second database (Base 2) that we used comes from a recent research. The study was conducted in the Central region of Cameroon, in the Okola district. It began in August 2015 and ended in January 2016.

**Figure 1:**
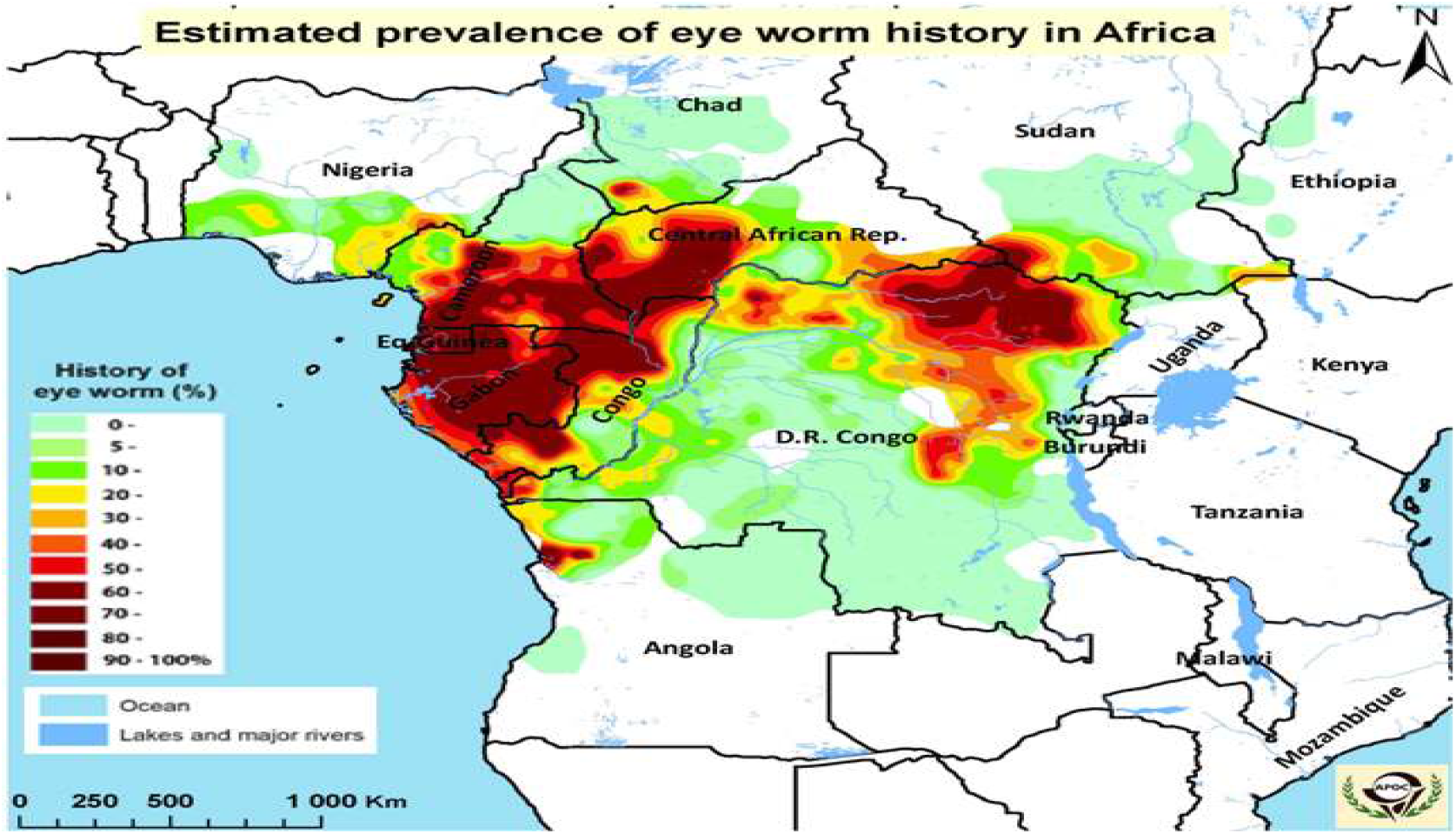
Distribution of loasis endemicity in Central Africa, from Zoure and al. (2011).

- For these data, it was carried out of measurement and reading.
- For the measurement, on each patient, two samples were made and for its samples the microfilarial load of Loa loa was measured.
- For the reading: Each measurement was read twice.

We performed a descriptive analysis of the data to explain the variation in Loa loa load as a function of gender and residence as categorical variables. We calculated the minimum sample size using formulas that associate with hierarchical data, see Zadeh (2003) and Usami (2014). In addition, we used the R software, R Core Team (2021), to perform all analysis. For the implementation of regression analysis on discrete qualitative data, we draw on Zeileis and al. (2008).

## 2 Modeling using hierarchical linear mixed model

For modeling the microfilarial load, we used the Generalized Linear Mixed Model (GLMM) with the negative binomial distribution (*NB*), see Hilbe (2011) and Jourdan & Kokonendji (2002) as the distribution of the variable of interest. The Hosmer-Lemeshow test is used for model validation. The following notations have been adopted. The variable of interest is the Loa Loa microfilar charge and is denoted *Y* = Ch.Micro.

### 2.1 Notations and meanings

For the sake of simplicity, we use the following notations. A bold notation is a vector.

- *n*_*i*_ is the number of reads performed on individual *i* and *m*_*i*_ is the number of measure performed on individual *i*.
- *Y*_*il*_, with *i* = 1, …, *n* and *l* = 1, …, *n*_*i*_ is the response variable of the variable *Y*, reading *l* of the microfilarial load on individual *i* ;
- *Y*_*im*_, where *i* = 1, …, *n* and *m* = 1, …, *m*_*i*_ is the response variable of variable *Y*, measure *l* of the microfilar load on individual *i* ;
- 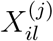, the *j*th variable observed on the *i*th individual with reading *l* ;
- ***u*** _*i*_ = (*u*_*i*1_, …, *u*_*iq*_)^*T*^, the vector representing the random effect on individual *i* ;
- The variable Sex is a categorical variable with two categories : male and female. We adopt the following notations:
- *x*_*i*1_, the variable noted `Age` ; *x*_*i*2_, the variable noted `Sex`; *x*_*i*3_, the variable noted `place of residence` of individual for *i* = 1, …, *n*;
- ***x*** _*i*_ = (1, *x*_*i*1_, *x*_*i*2_, *x*_*i*3_)^*T*^.

The data (individuals and measures or readings) suggest a hierarchical linear mixed model.

### 2.2 Presentation of the assumptions of the hierarchical linear mixed model

For the achievement of the objectives, we postulate a linear mixed model, see Martinez (2006) with the exception of the normality assumption and whose assumptions are as follows.

**The first assumption of the model** For an individual *i*, conditionally to the random effects ***u*** _*i*_, the *Y*_*il*_, *l* = 1, 2, …, *n*_*i*_ (respectively *Y*_*im*_, *m* = 1, 2, …, *m*_*i*_) are independent and follow a negative binomial distribution of parameters *α*_*i*_ and *δ*_*i*_, and we note :

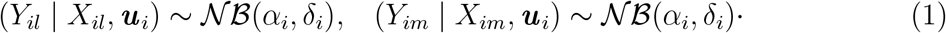

The density distribution of negative binomial of parameters *α* and *β* is given by

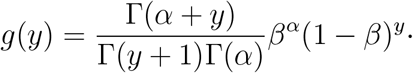

The microfilar load, generally follows a Poisson law of parameter which varies according to the population in which we are, following a gamma law. This gamma-Poisson mixture gives a negative binomial distribution. We have then, according to (1), the expectation and the variance of the conditional distribution are given by :

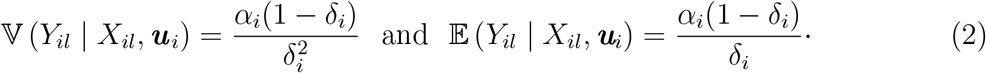

In the following, we will work with the notation *Y*_*il*_ and by analogy to *Y*_*im*_.

**The second hypothesis: Variance-covariance matrix and random effects**.

- The random effects ***u*** _*i*_ are assumed to follow a multivariate normal distribution with diagonal matrix *D*_0_.
- We assume that *δ*_*i*_ = *δ, ∀i* = 1, …, *n* to simplify the model.

**The third hypothesis: Link function** For *p* and *q* non-zero natural numbers such that *q ≤ p*, we suppose that for an individual *i* we have:

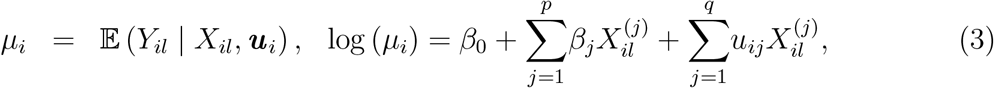

where

- *β*_0_ is the logarithm of the mean of the readings of all individuals ;
- *β*_1_, *β*_2_, …, *β*_*p*_, the fixed effects parameters ;
- 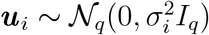, *I*_*q*_ is the identity matrix of order *q*.

Using the equation (2) and (3), we obtain *δ*_*i*_ = *α*_*i*_*/*(*α*_*i*_ + *µ*_*i*_). It follows that *∀ l* = 1, …, *n*_*i*_:

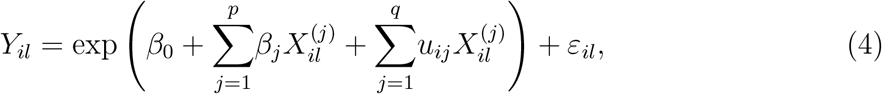

*ε*_*il*_ represents the error on the reading number *l* of the individual *i*. The final model is chosen using the AIC criterion.

### 2.3 Method of estimating the hierarchical linear mixed model parameters

The maximum likelihood or restricted maximum likelihood estimator has, under regularity conditions concerning the likelihood function, properties of almost certain convergence and asymptotic normality. For ***u*** = (***u*** _1_, …, ***u*** _*n*_) and ***y*** = (***y*** _1_, …, ***y*** _*n*_) et ***y*** _*i*_ = (*y*_*i*1_, …, *y*_*ip*_)^*T*^, where *p* = 3, the expression of the conditional likelihood for the model is

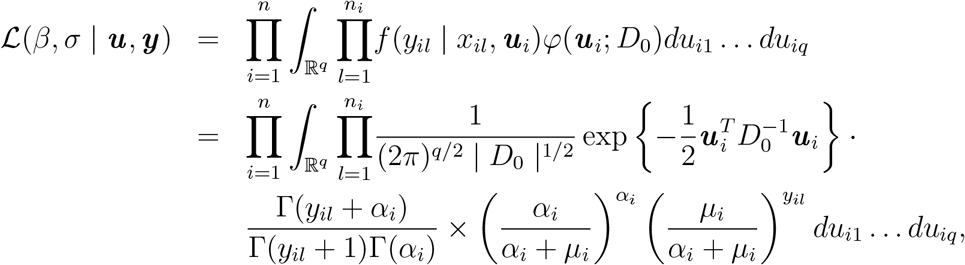

because (*Y*_*il*_ | *x*_*il*_, ***u*** _*i*_) *∼* 𝒩 ℬ (*α*_*i*_, *α*_*i*_*/*(*α*_*i*_+*µ*_*i*_)) where *µ*_*i*_ = ***x*** _*i*_*β* +***z*** _*i*_***u*** _*i*_ and ***u*** _*i*_ = (*u*_*i*1_, …, *u*_*iq*_)^*T*^, *β* = (*β*_0_, …, *β*_*p*_)^*T*^ et *σ* = (*σ*_0_, …, *σ*_*q*_)^*T*^. Consequently, the log-likelihood is given by :

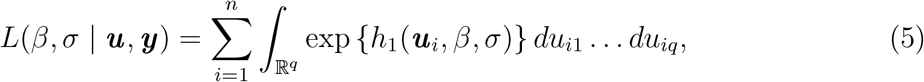

where

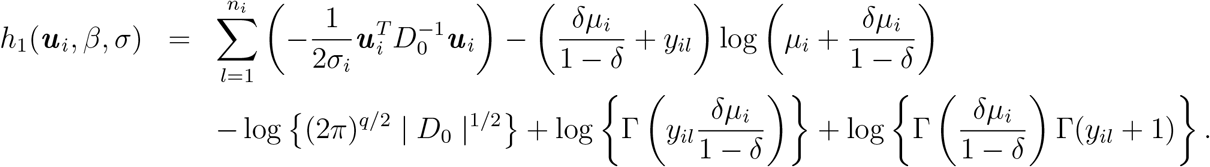

The complexity of this integration does not allow to calculate it directly. We then use numerical approximation methods or other methods that can get around this difficulty.

### 2.4 Calculation of measurement and reading uncertainties

Given the very limited number of samples and readings per individual, we will group the last two levels (sampling, reading) into a single level that we will call measurement. We recall the link between the variance and the mean by :

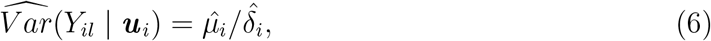

will allow to take into account simultaneously the variability related to the sampling and the variability related to the measurement. Thus, as in the first case above, we can estimate the uncertainty, see Salicone (2007) and Willink (2013), of measurement by 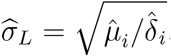 The same formulas apply for the calculation of uncertainty of reading the microfilar charge of Loa loa.

## 3 Results of data analysis

In this section, we present the proposed modeling results for estimating the measurement and reading uncertainty. All the results obtained in this article come from programming with the R software, see R Core Team (2021).

### 3.1 Descriptive analysis

As a first step, we need to look for the distribution of the variable of interest which is the Loa loa microfilarial load. We present the validation of the hypothesis of negative binomial distribution associated the microfilarial load of Loa loa.

For these data, it was carried out of measurement and reading, a study of adequacy by the test of Chi2 of Pearson. We tested in a first time, the test of Adequacy to the Poisson law, see Mizère and al. (2006) which failed.

**Table 1:**
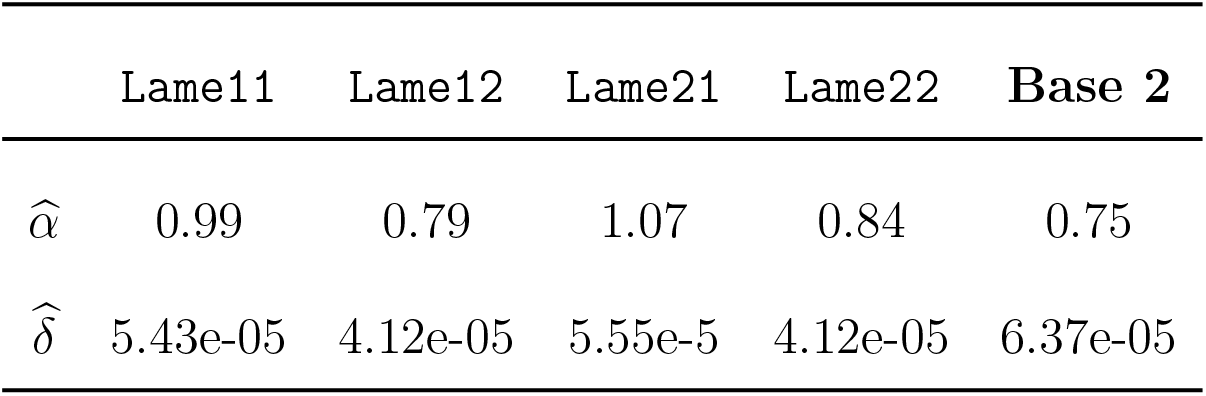
Estimated values of the parameters of the **𝒩ℬ**(negative binomial distribution) of the data of Base 1 and Base 2.

|  | Lame11 | Lame12 | Lame21 | Lame22 | Base 2 |
| --- | --- | --- | --- | --- | --- |
| $\hat{\alpha}$ | 0.99 | 0.79 | 1.07 | 0.84 | 0.75 |
| $\hat{\delta}$ | 5.43e-05 | 4.12e-05 | 5.55e-5 | 4.12e-05 | 6.37e-05 |

**Figure 2:**
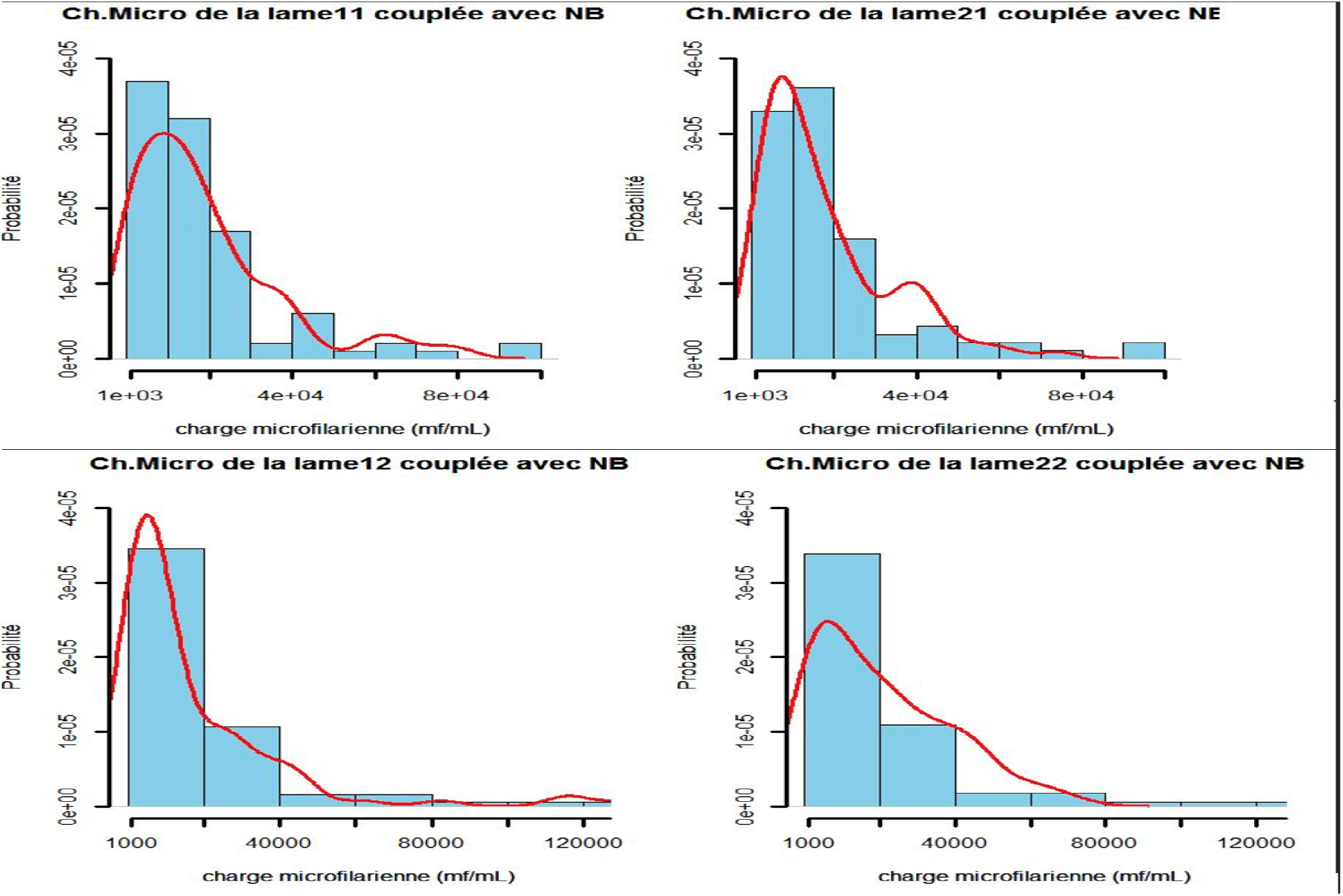
Adaptation of the negative binomial law of parameters (*a, λ*) to the microfilar charge of Loa loa whose parameters are on the table 1 as a function of the blade

The results allow us to conclude that the microfilar charge of Loa loa follows a negative binomial distribution as stated in the first hypothesis.

**Table 2:** Values of the *χ*^2^ statistic with the value of 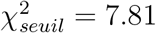.

| Lame | Lame22 | Lame22 | Lame21 | Lame22 | Tableau2 |
| --- | --- | --- | --- | --- | --- |
| value $\chi^2$ observed | 0.93 | 0.62 | 5.16 | 5.46 | 2.11 |

### 3.2 Parameter estimation results of the linear mixed model

The parameter estimates for the model loglikelihood function equations (5) is given in Tables 3 and 4 respectively. The AIC values presented use the formulas from the article Anderson & Burnham (1994) considering the fact that we note an overdispersion and the negative binomial distribution fitting.

**Table 3:** Estimated model parameters on reading and the other coefficients.

| **Model :** *glmer.nb(formula = Ch.Micro* |  |  |  |  |
| --- | --- | --- | --- | --- |
| *Lieu.residence+ ~(Lieu.residence—tnt.id)* |  |  |  |  |
| Min | 1Q | Median | 3Q | Max |
| -3.3552 | -0.4652 | 0.0009 | 0.4706 | 3.2429 |
| Parameters | Estimation | se | sd | p-value |
| $\beta_0$ | 8.86 | 0.062 | - | $< 2e - 16$ |
| $\sigma_0^2$ | 1.032 | | 1.01 | - |
| $\sigma_3^2$ | $9e - 3$ | - | 0.098 | - |
| AIC=10260.6; BIC=10282.4 ; $\delta = 540.34$ | | | | |

**Table 4:** Estimated model parameters on the measurement and the other coefficients.

| **Model : *glmer.nb(formula = Ch.Micro***<br>**Resid+(Lieu.residence—tnt.id)** |  |  |  |  |
| --- | --- | --- | --- | --- |
| Parameters | Estimation | se | sd | p-valeur |
| $\beta_0$ | 8.86 | 0.062 | - | $< 2e - 16$ |
| $\beta_1$ | -1.26 | 1.8e-1 | - | $9.52e - 12$ |
| $\sigma_0^2$ | 0.493 | | 0.7 | - |
| $\sigma_3^2$ | 0.358 | - | 0.6 | - |
| AIC=7426.4; BIC=7450 ; $\delta = 18.25$ | | | | |

The equation of model is

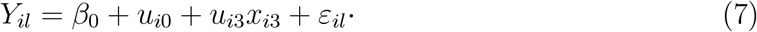

In the case where we estimate the parameters for the measurement uncertainty, we present the results in the table 4.

The equation of model is

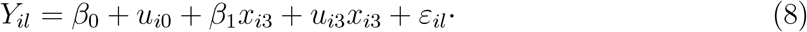

We conduct the Homser-Lemeshow test to confirm the selected models. The results of this test are summarized in the Table and we validate that the method of selection of the models of Hurvich & Tsai (1995).

In view of the results, with significant values, we retained these models. The uncertainty values summarized in the whole sample are estimated.

**Table 5:** Summary of the Hosmer-Lemeshow tests of validation of the model of *Ch*.*Micro*

| Model | Statistic value | P-value |
| --- | --- | --- |
| Model Read L | 15.2 | 0.2 |
| Model Measure | 28.37 | 0.57 |

**Table 6:** Summary of the uncertainties and Pari Interval of `textitCh.Micro`.

| Ch.Micro | Reading Uncertainty | Pari Interval |
| --- | --- | --- |
| 8,000mf/mL | 3.84 | 6,723.15-11,264 |
| 30,000mf/mL | 7.45 | 26,819.55-35,152.09 |

**Table 7:**
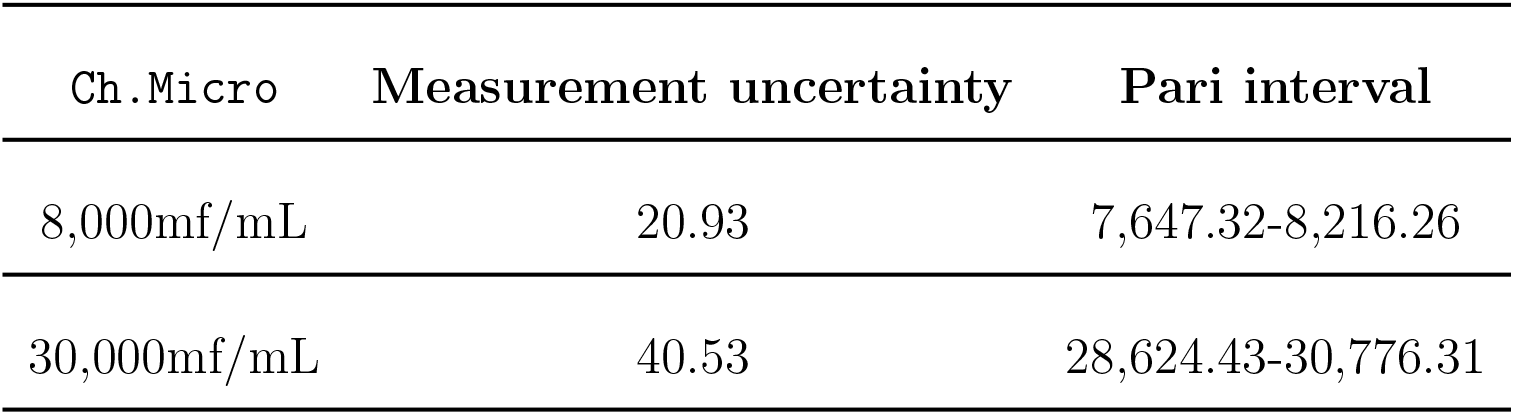
Summary of Uncertainties and Pari Intervals(PI) from *Ch*.*Micro*.

## Discussion

For the control of lymphatic filariasis, extensive population-based research has identified endemic areas. WHO recommends that the drug be administered to individuals with microfilarial loads of less than 30,000 mf/ml in areas where loasis and lymphatic filariasis are co-endemic. Examinations to assess the microfilaria count are very important in assessing the degree of microfilaremia. Measuring devices, no matter how good they are, can significantly influence the microfilarial load. Our study focused on the variation in the measurement and reading of the microfilarial load of Loa loa. This study allowed us to estimate the uncertainty of both the reading and the measurement of the microfilar load of Loa loa. During the realization of this work, we were confronted with difficulties. Its difficulties were manifested by the fact that the conclusions and results of our study have limitations. Among these, we can mention:

- In modeling Loa loa’s microfilarial load, we did not take into account the person who took the reading (measurement error related to the person measuring) ;
- In our work to explain the variability of the measurement of the Ch.Micro, there may be other variables to measure on individuals (height, etc) contributing to a good explanation of the sources of variability.

We also note the evolution of techniques or methods of assessing microfilaremia that allow for error mitigation, see Mouri and al. (2019).

## Conclusion

The fight against certain endemic diseases, in particular loasis, in third world countries is still problematic due to several factors: whether it is the management, the hygiene rules, etc., or the laboratory capable of doing in-depth research for these diseases, much remains to be done. It is in this context that some research centers have been looking for information to provide solutions to these ills that hinder development. It is in one of these researches that the CRFilMT, in collaboration with the Ministry of Health in order to fight against lymphatic filariasis, is confronted with the problem of the persistence of the disease due to the presence of the *loase*. Indeed, these researches are confronted with major difficulties in particular what was the object of our study in this thesis: Estimate the uncertainty of counting the microfilarial load of Loa loa. We have tried to answer this question by using statistical tools that are deemed necessary to give a precise idea about this quantity. In a first step, we estimated the uncertainty reading of the Loa loa microfilarial load and then in a second step, the uncertainty of measurement of the Loa loa microfilarial load. This work also allowed us to say that the uncertainty of measurement or reading of the microfilar load depends on the microfilar load counted in the individual, that is to say as the microfilar density increases in an individual, the uncertainty also increases.

At the conclusion of this work, we strongly recommend :

- the authorities at various levels, in charge of the coordination of the activities of fight against the neglected tropical diseases, more precisely the loasis, to put at the disposal of the laboratory technicians, apparatuses having a good precision;
- the various investigators and other stakeholders in the fight against lymphatic filariasis to pay particular attention to the enumeration of the microfilarial load, taking into account.

## Data Availability

All data produced in the present study are available upon reasonable request to the authors

## Acknowledgements

The author would like to thank the CRFilMT for providing data for the completion and writing of this paper. Special thanks to the Director, Prof. Tchatchueng, who accepted that I work with him.

## Author contributions

Talagbé Gabin Akpo is the person who followed the steps of design, intellectual content definition, literature search, data analysis, statistical analysis, manuscript preparation and manuscript editing.

